# Psychosocial impact of the Covid-19 pandemic: Identification of most vulnerable populations

**DOI:** 10.1101/2021.03.20.21254029

**Authors:** Judith Farrés, José Luis Ruiz, José Manuel Mas, Lilibeth Arias, Maria-Rosa Sarrias, Carolina Armengol, Pere-Joan Cardona, José A. Muñoz-Moreno, Miriam Vilaplana, Belén Arranz, Judith Usall, Antoni Serrano-Blanco, Cristina Vilaplana

**Affiliations:** Anaxomics Biotech SL. C/ Diputació, 237, 1r 1a, 08007 Barcelona.; Experimental Tuberculosis Unit (UTE). Fundació Institut Germans Trias i Pujol (IGTP). Universitat Autònoma de Barcelona (UAB). Edifici Mar. Can Ruti Campus. Ctra. de Can Ruti, camí de les Escoles, s/n, 08916 Badalona, Catalunya, Espanya.; Centro de Investigación Biomédica en Red de Enfermedades Respiratorias (CIBERES). Av. Monforte de Lemos, 3-5, pavelló 11, planta 0, 28029, Madrid, Espanya.; Innate Immunity Group. Fundació Institut Germans Trias i Pujol (IGTP). Can Ruti Campus. Edifici Muntanya. Ctra. de Can Ruti, camí de les Escoles, s/n, 08916 Badalona, Catalunya, Espanya.; Centro de Investigación Biomédica en Red de Enfermedades Hepáticas y Digestivas (CIBEREhD). Av. Monforte de Lemos, 3-5, pavelló 11, planta 0, 28029 Madrid, Espanya.; Childhood Liver Oncology Group, Germans Trias I Pujol Research Institute (IGTP), Program of Predictive and Personalized Medicine of Cancer (PMPCC), Badalona, Spain.; Fundació Lluita contra la SIDA (FLS) - Servei de Malalties Infeccioses, Hospital Germans Trias, Badalona.; Facultat de Psicologia i Ciències de l’Educació, Universitat Oberta de Catalunya (UOC), Barcelona.; Parc Sanitari Sant Joan de Déu. C/Antoni Pujadas, 42, 08830 Sant Boi de Llobregat, Catalunya, Espanya.; Centro de Investigación Biomédica en Red de Salud Mental (CIBERSAM). Av. Monforte de Lemos, 3-5, pavelló 11, planta 0, 28029 Madrid, Espanya.; Institut de Recerca Sant Joan de Déu. C/ Santa Rosa, 39-57, 3a planta, 08940 Esplugues de Llobregat, Catalunya, Espanya.; Centro de Investigación Biomédica en Red de Epidemiología y Salud Pública (CIBERESP). Av. Monforte de Lemos, 3-5, 28029 Madrid, Espanya.

## Abstract

With the aim of analyzing the psychosocial impact of Covid-19 pandemic on society in general and health care workers in particular, we developed a 74-question survey questionnaire which was shared through social media. After analyzing 56,656 responses obtained during the first pandemic peak, the results showed an early and important negative impact on family finances, fear of working with Covid-19 patients and ethical issues related to Covid-19 care among healthcare workers (HCW). We have identified 7 target groups at higher risk of impaired mental health and susceptible to benefiting from an intervention: women, under 42 years of age, people with care burden, socio-economically deprived groups, people with unskilled or unqualified jobs, Covid-19 patients, and HCW working with Covid-19 patients. These results should encourage the active implementation of specific strategies to increase resilience in these groups and to prepare an adequate organizational response.

**Summary box:** *What is already known?:*

- Studies in small cohorts have reported an important impact of the Covid-19 pandemic on the general population at several levels
- According to previous studies in small cohorts, approximately 20% of the population suffered from impaired mental health status due to the pandemic

*What are the new findings?:*

- We have studied 56,656 survey questionnaires to assess the impact of the Covid-19 outbreak on health status, family finances, habits, general health and mental health status, and ethics, especially in health care workers
- We have identified 7 target groups susceptible to benefitting from an intervention, and which should be taken into account when designing new contention measures against the pandemic

*What do the new findings imply?:*

- The design and active implementation of interventions to build individual resilience, especially for the targeted populations described, and preparation of an appropriate organizational response are key
- The results obtained in this project could help local and national Governments to design or adjust coping measures against future outbreaks

## 1. Introduction

On 30 March 2020, 78,797 confirmed cases of SARS-CoV-2, 6,528 deaths and 14,709 patients who had recovered were reported in Spain [1]; 16,157 cases and 1,410 deaths were recorded in Catalonia [2]. Case fatality (8%) was calculated for the registered cases, although the mortality rate was uncertain and the total number of cases (including those undiagnosed and with mild symptoms) were unknown. At that time, there was local transmission of SARS-CoV-2 in the community. Everyone with a compatible respiratory condition was considered likely to be a case of SARS-CoV-2 although the etiological diagnosis could not be made for all suspected cases in the context of a health emergency because of the lack of kits and the saturation of the health system [3,4].

Other major outbreaks of infectious diseases such as Ebola have demonstrated that there is an important impact on individuals and communities. The psychological effects of the disease itself as well as the traumatic experiences of loved ones are seen at individual level. At community level, health services, social systems and economic productivity are severely affected [5].

Two months after the first case reported in Spain and 2.5 weeks into the quarantine and self-isolation of the region of Catalonia, the emotional burden of the general community had increased. An important impact on mental health and emotional burden by SARS-CoV-2 epidemics and mass quarantines which have been implemented in other epidemics context has been reported [6–9]. Moreover, because a certain level of anxiety is necessary for the adoption of recommended precautionary measures against infection outbreaks [10], and for the successful implementation of public health interventions, a better understanding of people’s attitudes and the assessment of psychological impact on them should be mandatory.

On the other hand, 2,600 (16%) of the confirmed cases in our setting by March 30^th^ 2020 affected healthcare workers (HCW). Besides their obvious increased risk of being infected, the HCW facing the SARS-CoV-2 epidemics on the frontline (emergency rooms, ICUs, and other depts.) were put under high levels of stress and anxiety. This worsened as the tension in the Health Systems increased, requiring them to face important ethical dilemmas including triage of patients. Additionally, the SARS epidemic proved that frontline healthcare workers not only suffered from chronic stress at the time, but that this lasted for at least one year after the epidemic wave was over [11].

In the face of all this, we decided to evaluate the impact of the first wave of the Covid-19 pandemic on both the general population and HCW, specifically on their socio-economic status and their psychological distress.

## 2. M&M

### 2.1. Ethics

The study was reviewed and approved by the corresponding Ethics Committee, the Comitè Ètic de l’Hospital Universitari Germans Trias i Pujol; and conformed to the principles embodied in the Declaration of Helsinki. The ethical clearance was obtained before starting the project. The survey was created and shared complying with the European General Data Protection Regulation (GDPR), and all data was processed anonymously. The project is registered in ClinicalTrials.gov under the identifier NCT04378452.

### 2.2. Study procedures

An anonymous online survey was created with the Typeform software (Typeform SL, Barcelona, Spain). It included 74 questions on demographic data (12 questions), socio-economic sphere (8 questions), habits and health status related to Covid-19 during confinement (13 questions) and mental health dimension (through questions related to depression, anxiety, stress and Post-Traumatic Stress Disorder [PTSD] symptoms [41 questions]) (Supplementary Table 1). The survey was shared in 5 different languages (Catalan, Spanish, English, Italian, and French) through social media using snowball sampling. The data were downloaded as a spreadsheet file (Excel Microsoft Office) after collection and deleted from the Typeform software.

### 2.3. Analysis and Statistics

Since there were no specific criteria for age stratification or the population density (inhabitants / km2) of the municipality where the respondents lived that was significant for all questions, it was decided to divide these categories in the cohort into groups containing a similar sample size. Thus, and taking into account the volume of responses obtained, age ranges have been determined statistically so that they are homogeneous in terms of number of surveys completed by group.

The questions were grouped into indexes (socioeconomic precariousness index, depression index, anxiety index, stress index, or PTSD). The scores of the socio-economic precariousness index and population density by the respondents were segmented into 4 groups each. The criteria for segmentation were established in order to obtain balanced groups in terms of the number of respondents in each category.

We determined 4 ranges of age: <42 years old, 42-52, 52-61 and >61 y.o. The 4 score ranges of the 0-19 scale of socio-economic precariousness established were: low precariousness ≤7 points, mid-low=7-8.5, mid-high=8.5-10 and high >10 points.

All results were obtained taking into account the fact that the respondents were part of the totality of the cohort of respondents. Responses were also analyzed in total by category and broken down into percentages according to conditional distributions taking into account; on the one hand the gender of the respondents, and on the other their age group.

We took the non-binary gender and those who preferred not to say which gender they identify as into account when analyzing the results, as this enriches the conclusions. However, statistical analysis, often does not take into account the minimum volumes of responses and therefore only the groups of women and men were compared.

Response percentages were calculated based on the number of respondents for each answer out of the total number of responses to each question. To assess whether the categorical variables were significantly related or not, we applied the Chi-Square test independently in the observed counts. We conducted a bivariate analysis between scores and sociodemographic variables. Differences in score distribution between different groups were assessed by comparing probability distributions using a two-band Wilcoxon-signed rank test and collecting the p-value using Matlab’s ‘signrank’ function [12,13].

All tests were applied bilaterally using a significance of 5% (p <0.05).

## 3. Results

The complete dataset results generated is available at: https://zenodo.org/badge/DOI/10.5281/zenodo.4608502.svg

### 3.1. Characteristics of the cohort

We analyzed 56,656 questionnaires. The characteristics of the cohort are described in Table 1. Differences between categories by gender and age are described in Supplementary Table 2. The majority of respondents were females (70.4%), and from Catalonia. Those living most precariously were under 42 years old, with 18.43% sharing an apartment/house. (p<0.01). Most respondents had a degree (42.62%), and a qualified job (36.13%). 9% of total respondents worked in the healthcare sector. Most unemployed people were in the younger age range (7.6%) and in the non-binary/those who preferred not to say groups (approximately 12% each). Up to 60% of the total declared that they were taking care of someone: 24.81% caring for children of <16 years and 15.11% caring for parents. Women were caregivers more frequently than men (p<0.01). The burden of care was also higher for women and people of 42-61 years old (p<0.01) and concerningly high for 4.79% of total respondents.

**Table 1:**
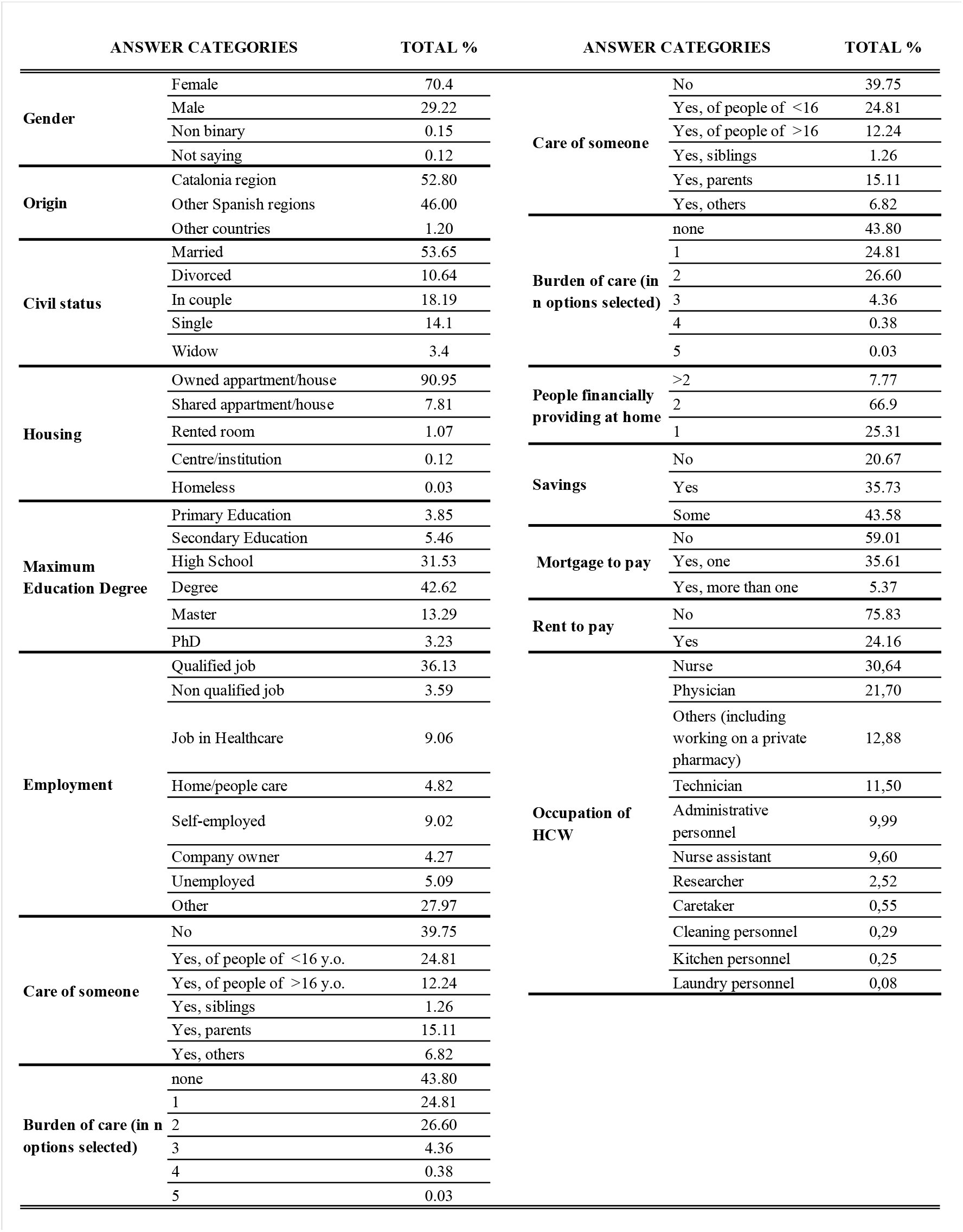
Characteristics of the cohort.

### 3.2. Impact of the pandemic on the General population

The impact on general population according to the responses obtained to the questionnaire is described in Table 2. Categories of responses by gender and group are described in Supplementary Table 2. 85.32% of the cohort declared they were remaining at home. Those working in essential services were mostly women or of non-binary gender, and the percentage of women was also higher amongst those who were obliged to go to work on-site (p<0.01).

**Table 2:**
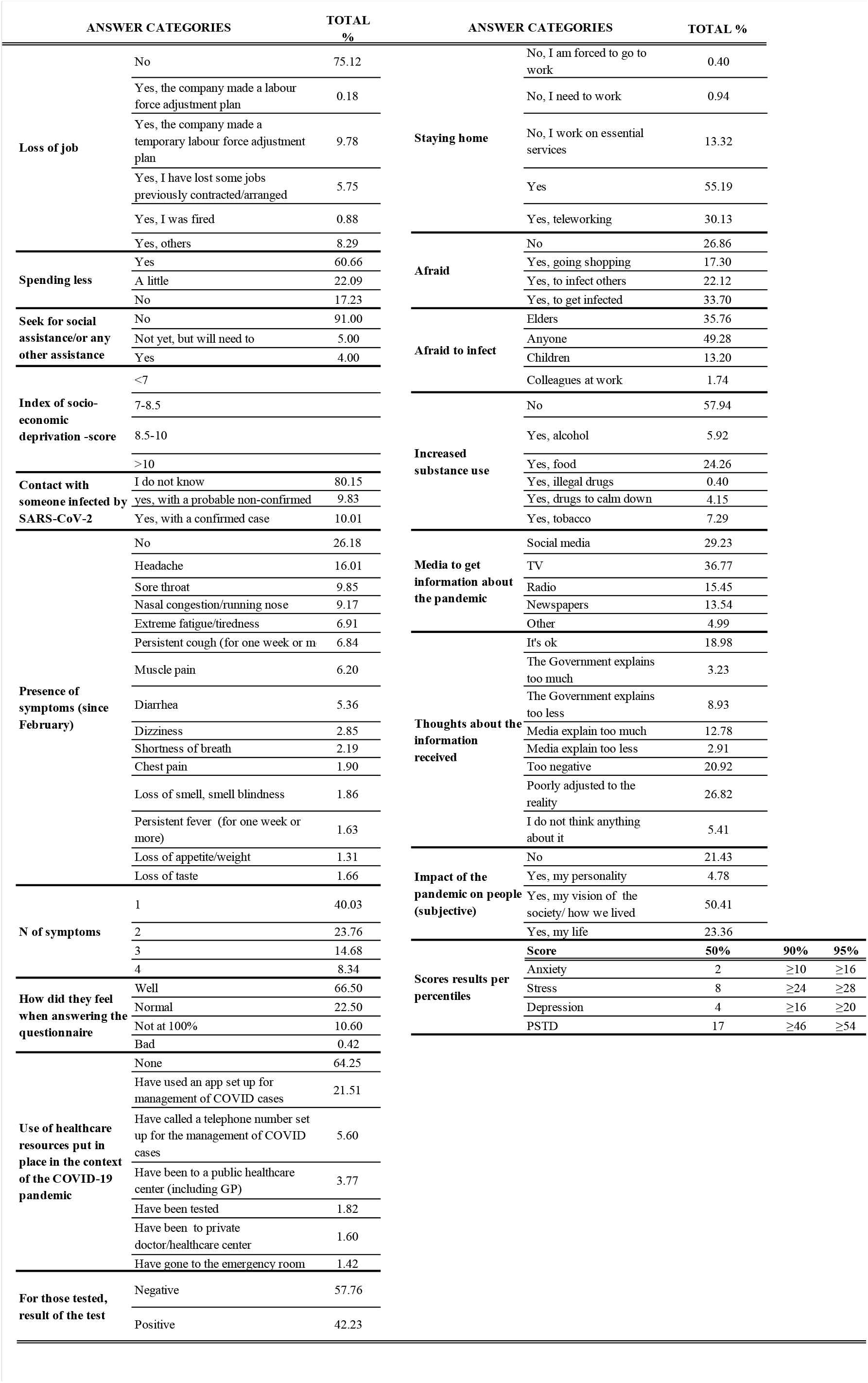
Impact of the pandemic on the General population.

Only 2 weeks after starting the lock-down, 25% of the cohort had already lost their job or work. People under 52, as opposed to people over 52, and men, as opposed to women, were the most affected (p<0.01). 20.67% of the respondents declared that they had no savings at all (Table 1). After the start of measures announced by the authorities to cope with the pandemic, 82.75% of respondents declared that they were being careful or had decreased their expenses. Up to 8.78% of respondents declared that they had used social services help or that would need to use it soon. Those under 52 and people identifying as non-binary gender or preferring not to say were the most affected (p<0.01 and p<0.05, respectively). Those under 42 years, followed by people over 61 and people identifying as non-binary gender were the ones who showed higher precariousness index values (p<0.01).

The 19.84% of respondents declared that they had had contact with someone infected by SARS-CoV-2, half of them with a confirmed or probable case and this was more frequent for women under 52 (p<0.01). 35.75% declared that during the previous 14 days they had used at least one existing healthcare resource or one put in place by the authorities in the context of the pandemic, and 64.25%, had used none. 73.82% declared to have had one or more symptoms compatible with Covid-19. The most frequent symptoms were headache (16.01%), sore throat and nasal congestion (9.85% and 9.17% respectively). Only 1.76% of people with one symptom or more had received a PCR test and only 1.81% of those declaring three symptoms or more. Women and under 42 said that they felt worse at the moment they answered the survey than people in other groups (p<0.01).

The 42.05% of respondents said they had increased their consumption habits: in most cases of food. Women under 42 showed the largest increase in consumption, except for illegal drugs, compared with other groups (p<0.01).

Most people said TV was their source of information on the pandemic (36.77%), followed by social media (29.23%). 30% of people only used one source, 37.84% 2 sources and 23.05% used 3. There was no difference across gender or age groups. 26.82% declared that the information given did not accurately reflect reality (more frequent in women and people over 52 (p<0.01), and another 20.92% said that it was too negative or too sensationalist (more frequent in men and people under 42 (p<0.01). 73.13% declared that they were afraid or worried, these including more women, but a lower percentage of people over 61 (p<0.01).

The 78.56% of the cohort declared that the pandemic had changed them, most of them (50,41%) in the way that they see society/how we used to live. Those most affected were women (more than men) and those under 42 vs the >61 (p<0.01 in both cases).

### 3.3. Impact of pandemic on HCW

A total of 5,104 people (9.05% of the total) identified themselves as workers in the healthcare sector, most of them women. While the proportion women/men in the total cohort is 70/30 in this subgroup the proportion is 85/15. The impact on this population is detailed in Table 3. 41.65% of healthcare personnel declared that they had worked directly with Covid-19 patients, 32% of them while on duty. The majority of healthcare workers said that they were afraid to work with Covid-19 patients (75.87%). As it was a multiple-choice question, we know that around the 42.90% were afraid of transmitting the infection to their relatives/friends, 17.07% feared getting infected or transmitting it to other patients, and 4.28% were afraid of dying. Surprisingly, fear of dying decreased with age. In all cases it was higher percentages of younger HCW who said they were afraid (p<0.01).

**Table 3:** Impact of the pandemic on the General population

| ANSWER CATEGORIES |  | TOTAL % | ANSWER CATEGORIES |  | TOTAL % |
| --- | --- | --- | --- | --- | --- |
| **Having worked directly with COVID-19 patients** | No | 58.34 | **Ethical concerns** | No | 56.29 |
|  | Yes | 41.65 |  | No, I follow protocols | 25.09 |
| **Fear of working with COVID-19 patients** | No | 24.13 |  | Yes, with selection of patients and/or protocols for selection of patients or therapeutic indications | 9.41 |
|  | Yes | 75.87 |  | Yes, others | 9.19 |
| **Fear of working with COVID-19 patients** | No fear | 14.58 | **Problems faced by healthcare professionals, grouped** | Having worked without sufficient protection | 25.68 |
|  | Scared of transmitting the virus to other non-COVID patients | 14.95 |  | With patients triage or protocols for patients triage or therapeutic indication | 16.28 |
|  | Scared of transmitting the virus to own people (family, colleagues) | 42.90 |  | With the protocol for case management. | 11.46 |
|  | Scared of being obliged to take medical decisions representing an ethical dilemma for me (patient selection, application of protocols) | 6.26 |  | With the protocol for End-of-Life management | 8.94 |
|  | Scared of being infected | 17.01 |  | With institution management or orders from superiors. | 8.02 |
|  | Afraid of dying | 4.27 |  | With the disjunctive of having to/wanting to go to work at first line and not being able/wanting to do it. | 6.88 |
|  |  |  |  | With the prioritization of dispensing protective material (facial masks, EPIs) or tests. | 5.27 |
|  |  |  |  | With the impact of the outbreak and/or lockdown on some populations (chronic or mental health patients, elders, etc.) | 3.89 |
|  |  |  |  | Others (non-specified) | 3.89 |
|  |  |  |  | With problems due to the organizational changes. | 3.66 |
|  |  |  |  | With management of information given to patients/their families, and related problems (including confidentiality issues). | 3.44 |
|  |  |  |  | With colleagues attitudes | 2.52 |

More than 6 percent of healthcare workers (6.27%) were worried of taking medical decisions that represented an ethical problem for them. In fact, nearly 18.60% of them said that they had ethical problems/dilemmas/issues while working. Of these, the younger the respondents, the higher the percentage, especially with the patient triage and obligatory protocols (p<0.01). As many as 437 of 5,104 healthcare workers decided to explain to us which ethical problems they had had. We have grouped the problems and issues that the professionals listed, and the results are found in Table 3.

### 3.4. Impact of the pandemic on mental health status

Table 4 summarizes the conditions found statistically significantly associated (p<0.05) with the mental health symptoms evaluated. According to this table, we have identified 7 target groups susceptible to benefitting from an intervention, and which should be taken into account when designing new contention measures to cope with the pandemic: 1) women; 2) people under 42; 3) caregivers; 4) people working in essential services or non-qualified jobs; 5) people with a higher precariousness index; 6) Covid-19 patients and 7) healthcare personnel, especially those working with Covid-19 patients.

**Table 4:** Conditions statistically associated to the mental-health scores results.

| Factors: | Statistically association to: |  |  |  |  |  |  |
| --- | --- | --- | --- | --- | --- | --- | --- |
|  | Depression Index | Anxiety Index | Stress Index | PSTD Index | Evitation Index | Intrusion Index | Hyperarousal Index |
| Risk | p | p | p | p | p | p | p |
| Women | 0.019 | 0.003 |  | 0.000 | 0.007 | 0.034 | 0.027 |
| <42 y.o. |  | 0.008 |  |  |  |  |  |
| Caregivers |  | 0.002 | 0.039 | 0.006 |  | 0.050 |  |
| Adults with higher perception of the difficulty of quarantine for children and the whole family (score in a 10-points scale) vs 0 |  |  |  | 0.041 |  | 0.032 | 0.022 |
| Living in a middle-high density population town |  | 0.031 |  |  |  |  |  |
| Living in a shared apartment/house |  | 0.006 |  |  |  |  |  |
| Living in a rented room |  | 0.039 |  |  |  |  |  |
| Declaring to be homeless |  |  |  | 0.044 |  |  |  |
| High deprivation index (>10) |  | 0.015 |  |  |  |  |  |
| Going to work because job on essential services |  | 0.011 |  |  |  |  |  |
| Being a healthcare worker and to be afraid of attending COVID-19 patients | 0.017 |  |  |  | 0.023 |  |  |
| To have been in contact with a COVID-19 patient |  | 0.006 |  | 0.038 |  |  |  |
| Having had symptoms compatible with COVID-19 | 0.021 | 0.002 |  | 0.008 |  |  |  |
| Having used all healthcare resources put in place in the context of the COVID-19 pandemic |  |  | 0.039 | 8641,000 | 0.007 |  | 0.011 |
| To be afraid (of getting infected, to infect others, to go shopping) |  | 0.000 | 0.036 | 0.000 | 0.003 | 0.012 | 0.006 |
| To have increased the consume of at least one substance |  | 0.006 |  | 0.008 |  |  |  |
| To use 3 media to get information about COVID-19 |  | 0.033 |  |  |  |  |  |
| Protection | p | p | p | p | p | p | p |
| >61 y.o. |  | 0.006 |  | 0.05 |  |  |  |
| To be married |  | 0.007 |  |  |  |  |  |
| Being a widow |  |  |  | 0.020 | 0.011 |  |  |
| To have a qualified job |  | 0.008 |  |  |  |  |  |
| To have a PhD | 0.019 | 0.010 |  |  | 0.031 |  |  |
| Feeling well |  | 0.045 |  | 0.037 |  |  |  |

## 4. Discussion

Researchers have already sounded the alarm about how the Covid-19 pandemic may affect the mental health of the general population, and more specifically patients with previous physical or mental conditions (including previous mental disorders) [14,15] and people at risk due to their socio-economic conditions. The current study aimed to identify the impacts of the covid-19 pandemia at several levels using a questionnaire.

Our survey was disseminated through social media, thus the sample of population studied could not be controlled. However, this was a successful strategy to rapidly reach a large number of people in different settings and different regions, without exposing interviewers to infection. Even though this does not ensure representability, there is no other study that has reached such a huge number of subjects, as more than 50,000 completed questionnaires were obtained from geographical regions hit by the pandemic in different ways.

The criteria used to establish the age ranges, the population density and the socioeconomic precariousness index were statistical, in order to obtain balanced groups in terms of number of responses. This provides rigor but can be confusing because this segmentation is unusual and can lead to a certain bias.

As for the impact on the socioeconomic sphere, the highest level of precariousness, which according to what the results seem to reflect occurs in those under 42 years of age, is striking. Of particular concern is the fact that 25% of the people who responded to the survey in our study had decreased their workload due to the epidemic situation. According to the International Labor Organization (ILO), the reduction in employment is greater among women and younger and older people, who have all been particularly affected by the Covid-19 crisis. In our study, men are the ones who had lost more jobs or assignments previously contracted or hired, and we saw that higher percentages of those under 52 years old had been dismissed or submitted to a temporary labour force adjustment. Overall, global labour incomes have been estimated to have fallen by 10.7% during the first three quarters of 2020 (compared to the same period in 2019) [16], but we believe this could be much worse given our results. In addition, in our study, a quarter of respondents had no savings to deal with contingencies, and up to 8.78% stated that they had applied for social benefits or that they would do so soon. All of this is important because as we have demonstrated in our results, socioeconomic precariousness was revealed to be one of the factors associated with higher scores on mental health indices, and this is even more worrying given that the incidence of the epidemic was also more pronounced in the poorest neighborhoods, at least in Barcelona [17]. We would also like to mention that more studies should be carried out to analyze the socio-economic precariousness of the group of non-binary people, as we have seen trends that have not been statistically evident but would be worth confirming.

According to the literature, approximately 20% of the population affected seems consistent [7,18,19], even if in some cases higher percentages have been found [20,21]. According to our results, we have identified up to 7 target groups at higher risk of impaired mental health status and susceptible to benefitting from an intervention, and which should be taken into account when designing new contention measures against the pandemic. In our study we did found an association of worse symptoms scoring with the presence of symptoms compatible with Covid-19 or having used all the healthcare resources put in place. However, as a real intervention based on these assumptions would be very costly and logistically difficult, we do consider instead that the target group for an intervention should be confirmed Covid-19 patients.

Other studies have also shown that being female, young, and having unstable work or income to be significant correlators of psychological negative impact [20–23]. Women are especially vulnerable as they bear the heavier burden of childcare and care of the elderly, suffer gender violence and have more precarious jobs. This effect, which is generalized in society, is even more obvious if the female sex is combined with characteristics of vulnerable groups [24]. Sex and gender biases have been identified as linked to Covid-19 outbreaks. In many settings, women appear to be slightly more likely to be diagnosed with Covid-19, which may in part be due to the fact that women account for the majority of health care workers around the world. Moreover, several studies have highlighted that fact that health staff who are women, younger or parents of dependent children are more vulnerable to psychological distress [25]. We also know that crises exacerbate gender inequalities: gender-based violence increased during confinement [26]; women were doing 3-10 times more care work than men; women faced significant barriers to healthcare due to lack of autonomy over their own sexual and reproductive health, inadequate access to health services, and insufficient financial resources [27]. In this sense, it is anticipated that the Covid-19 crisis will trigger an economic recession which will disproportionately impact the income and employment of the most vulnerable, particularly women [28]. In our setting it was mostly women who were responsible for caring for others. Caregiver adults with higher perception of the difficulty of quarantine for children and the whole family suffered more psychological distress than the other groups. This was previously identified in a cohort of parents in Italy, showing that their individual perception was associated with their stress levels and a negative behavioural and emotional impact on their children. As this study points out, some of the causes for this could be the impact of the situation itself both on the adults and the children, plus the effects of the school closure together with the need for working from home with a lot of new inputs. It not only has a negative effect on the adults, but on the children both indirectly [29] and directly [30]. Schools provide not only education, but also counselling and promote and imply healthy habits (healthy diet, physical exercise, social interaction), that might not be continued at home [30].

On the other hand, people over 60 years old were the vast majority of the total number of deaths all over the world [31]. While their frailty and an increased risk of suffering Covid-19 if living in nursing homes or similar facilities is true/undeniable, the elderly are key in Mediterranean countries, such as ours, as they take care of grandchildren when their parents go to work, so to quarantine and isolate them can be very disturbing for the whole of society. Moreover, Covid-19 and the consequences of isolating the elderly can be devastating, not only for their mental health but also as it contributes to a greater risk of morbidity, and this can be even worse in the more disadvantaged populations [32,33]. In this perspective, older people had more difficulties than younger people in adjusting to lockdown and social distancing rules. On the other hand, older people have proved that they have more resilience than younger people in other outbreaks and major hazards [34], something that our results also support by showing that older people were less afraid of dying than younger ones. All seniors showed anxiety and depression issues in China, and results were worse for females [35]. A Spanish study reported that up to 25.6% of a sample of adults with a mean age of 65 had symptoms of depression and 32.1% symptoms of avoidant coping style, and that having a current or past history of mental disorders highly influenced this, while the main protective factor was the ability to enjoy free time [36]. However, we found that younger people coped worse than older people with the mental burden due to the Covid-19 pandemic and the measures dictated to combat it. Differences between younger and older adults in emotional responses and recovery have been previously described, and several reasons for it have been hypothesized, including the fact that the elderly have a higher sense of meaning of life and that for them perceiving time as finite determines their priorities in terms of goals and behaviours [37]. In the context of the Covid-19 outbreak’s first wave, others have reported an increased negative impact on younger people compared with the elderly. A study in France after 2 weeks of confinement reported sleep problems and increased consumption of sleeping pills, with both more frequent in people under 35 compared to older people[38]. Young adults already face life changes which are stressful and the pandemic has worsened this, even if one out of five young adults might have been better off because of being removed from external pressures such as work and education and/or to having more time for close relationships [39]. A nice study in Switzerland concluded that for this specific population the distress related to lifestyle disruptions and hopelessness was higher than the perceived virus-related health risk [39], which others have already shown to be was relatively low [40]. Shanahan et al also showed that a good group to be selected for intervention could be females, migrants and young adults with higher pre-pandemic emotional distress including social exclusion [39]. Another factor which has been related to distress is the decrease of physical and social activity due to lockdown and other restriction measures decreed by Governments, which had a negative impact on psychological wellbeing of individuals including the elderly [41], but especially on the group of adolescents and young adults [40,42].

A non-negligible proportion of our respondents were HCW, who in Europe are mostly women [43]. Besides their obvious increased risk of being infected [44], facing the SARS-CoV-2 epidemics at the frontline may have put them under a lot of pressure, increasing levels of anxiety and chronic stress (as they faced tremendous overwork and suboptimal working conditions), which can last to up to a year afterwards [11,45,46].

A study carried out in a cohort of 9,138 HCW showed that 45.7% were at risk of suffering from a mental disorder [47], and another, which included 5,450, showed that 8.4% had suicidal ideation and behaviour [48]. In our study, being a HCW has been revealed as a positive factor for impaired mental health, especially for those working with Covid-19 patients and afraid of infecting others, which has proved to have an impact on outcomes [49].

This becomes worse as the tension in health systems increases, as front-line professionals work in a complex environment given the ethical challenges of the Covid-19, eliciting different dimensions of ethical dilemmas related to the situation itself and the measures dictated by the Government [50]. The shortage of hospital beds — and especially ICU beds — was also an important problem, contributing to the case fatality rate and implying a triage of patients in order to preserve the beds for those with an increased potential to survive [51–53]. The management of end-of-life situations was particularly worrying, as banning the support of relatives at the bedside had a very disturbing impact on patients and their families, but also on HCW mental health, workload, challenges and professional outcomes [54]. According to our results, nearly 8 out of 10 HCW declared that they were afraid of working with COVID patients, especially because of infecting others. Being obliged to work with lack of appropriate or sufficient personal protective equipment was one of the most frequent complaints of HCW who shared their narratives on the ethical concerns they suffered. This low sense of security had been previously pointed out in a small HCW cohort in Spain [55], in nurses in Poland [56] and in Latin America [57]. We found differences between women and men in terms of the fear of transmitting the infection to others, and this could be related to women’s jobs implying more exposure (as is the case for nurses, that in our cohort were mostly women). In our study those working in essential services also had higher psychological distress and this could be for the same reason, the low sense of security, plus the fear of being at higher risk of contracting the infection.

The 6.27% declared that that their fear was of making medical decisions that represented an ethical problem for them (patient selection or application of protocols), and this percentage was higher in younger people.

In fact, in our sample, one in five of the HCW declared that they had had ethical problems during those first weeks of the peak of the first wave, which is in line with other studies [54,58]; and approximately half of these had to do with patient selection or patient triage protocols/therapeutic indications. In our opinion, this fact should also be explored more thoroughly and actively followed up to prevent health professionals from being put into similar situations in the future.

## 5. Conclusion

Our study represents a photograph of the impact of the Covid-19 outbreak on the general wellbeing of the population and HCW, which should open the door for the elaboration of strategy proposals with the full participation of institutional leaders who are in a position to adapt policy to the real needs of the people. Previous work in smaller, selected cohorts (seniors, youth, etc.) has described the significant impact of the pandemic in a number of areas, including mental health problems in 20% of the population. In this project we have studied 56,656 completed surveys and analyzed the effects of Covid-19 on family finances, habits and attitudes, general health and mental health, and the day to day of health professionals. We were able to confirm the results noted by other smaller studies and to identify up to seven populations likely to benefit from an intervention: women; those under 42 years old; caregivers; people in a situation of socio-economic precariousness; essential workers or those with unskilled jobs; Covid-19 patients, and HCW, especially those working with COVID-patients. This data should be used to design and implement interventions to increase the resilience of these identified groups, as well as to prepare an appropriate organizational response. In this sense, some authors have published specific strategies that could be used to alleviate this suffering, especially in terms of increasing the adaptability of caregivers by providing tools for recognizing risk factors for emotional distress and managing mental health hygiene, but also response actions by public and private organizations aimed at identifying the employees most at-risk and establishing active mitigating and corrective measures [52,54,59–62]. We think it would be worthwhile studying how to actively implement and adapt these measures to our environment, not only in the health field but also by extending them to the groups we have identified. The results obtained could help local and national Governments and Public Health Services to design or adjust coping measures in the face of potential future outbreaks or other major hazards that might be difficult for society.

## Supporting information

Survey Questionnaire

Results broken down by category and according to gender and age group

## Data Availability

All data generated within this project is publicly available at Zenodo.

https://zenodo.org/badge/DOI/10.5281/zenodo.4608502.svg

## 7. Acknowledgements

We thank the people that answered the survey questionnaire online, as well as Jérôme Nigou, Valeria Bergalli, Paolo Salieri, Chiara Bertoldini and Harvey Evans, who kindly volunteered to translate/edit the questionnaire and website information to French, Italian and English languages.

## References

1. Ministerio de Sanidad Servicios Sociales e Igualdad; Ministerio de Economía y Competitividad; Instituto de Salud Carlos III. Situación del COVID-19 en España. Available at: https://covid19.isciii.es/.

2. Catalunya D de S de la G de. Comunicat del Departament de Salut, 29 de març de 2020. Available at: http://canalsalut.gencat.cat/ca/salut-a-z/c/coronavirus-2019-ncov/nota-premsa/?id=384272.

3. CDC. Patients with COVID-19. CDC website. 2020. Available at: https://www.cdc.gov/coronavirus/2019-ncov/hcp/disposition-hospitalized-patients.html%0D%0A%0D%0A%0D%0A. Accessed 20 March 2020.

4. European Centre for Disease Prevention and Control. Novel coronavirus disease 2019 (COVID-19) pandemic : increased transmission in the EU / EEA and the UK – sixth update. 2020. Available at: https://www.ecdc.europa.eu/sites/default/files/documents/RRA-sixth-update-Outbreak-of-novel-coronavirus-disease-2019-COVID-19.pdf.

5. Van Bortel T, Basnayake A, Wurie F, et al. Psychosocial effects of an Ebola outbreak at individual, community and international levels. Bull World Health Organ 2016; 94:210–214.

6. Rubin GJ, Wessely S. The psychological effects of quarantining a city. BMJ 2020; 368:1–2. Available at: http://dx.doi.org/ doi:10.1136/bmj.m313.

7. Wang C, Pan R, Wan X, et al. Immediate psychological responses and associated factors during the initial stage of the 2019 coronavirus disease (COVID-19) epidemic among the general population in China. Int J Environ Res Public Health 2020; 17.

8. Sim K, Huak Chan Y, Chong PN, Chua HC, Wen Soon S. Psychosocial and coping responses within the community health care setting towards a national outbreak of an infectious disease. J Psychosom Res 2010; 68:195–202.

9. Brooks SK, Webster RK, Smith LE, et al. The psychological impact of quarantine and how to reduce it: rapid review of the evidence. Lancet 2020; 395:912–920. Available at: http://dx.doi.org/10.1016/S0140-6736(20)30460-8.

10. Leung GM, Lam TH, Ho LM, et al. The impact of community psychological responses on outbreak control for severe acute respiratory syndrome in Hong Kong. J Epidemiol Community Health 2003; 57:857–863.

11. McAlonan GM, Lee AM, Cheung V, et al. Immediate and sustained psychological impact of an emerging infectious disease outbreak on health care workers. Can J Psychiatry 2007; 52:241–247.

12. Gibbons JD, Chakraboti S. Nonparametric Statistical Inference. 5th ed. Boca Raton: Chapman & Hall/CRC Press, Taylor & Francis Group, 2011.

13. Hollander M, Wolfe D. Nonparametric Statistical Methods, 2nd Edition. 1999.

14. Yao H, Chen JH, Xu YF. Patients with mental health disorders in the COVID-19 epidemic. The Lancet Psychiatry 2020; 7:e21. Available at: http://dx.doi.org/10.1016/S2215-0366(20)30090-0.

15. Cullen W, Gulati G, Kelly BD. Mental health in the COVID-19 pandemic. QJM 2020; 113:311–312.

16. International Labour Organization. ILO Monitor: COVID-19 and the world of work. Second edition. Updated estimates and analysis. 2020. Available at: https://www.ilo.org/wcmsp5/groups/public/@dgreports/@dcomm/documents/bri efingnote/wcms_740877.pdf.

17. Marí-Dell’olmo M, Gotsens M, Pasarín MI, et al. Socioeconomic inequalities in COVID-19 in a European urban area: Two waves, two patterns. Int J Environ Res Public Health 2021;

18. González-Sanguino C, Ausín B, Castellanos MÁ, et al. Mental health consequences during the initial stage of the 2020 Coronavirus pandemic (COVID-19) in Spain. Brain Behav Immun 2020;

19. Wang C, Pan R, Wan X, et al. A longitudinal study on the mental health of general population during the COVID-19 epidemic in China. Brain Behav Immun 2020;

20. Rodríguez-Rey R, Garrido-Hernansaiz H, Collado S. Psychological Impact and Associated Factors During the Initial Stage of the Coronavirus (COVID-19) Pandemic Among the General Population in Spain. Front Psychol 2020;

21. Jacques-Avinõ C, López-Jiménez T, Medina-Perucha L, et al. Gender-based approach on the social impact and mental health in Spain during COVID-19 lockdown: A cross-sectional study. BMJ Open 2020;

22. Ramírez Lpg, Arriaga RJM, Hernández-Gonzalez Ma, De la Roca-Chiapas Jm. Psychological distress and signs of post-traumatic stress in response to the COVID-19 health emergency in a Mexican sample. Psychol Res Behav Manag 2020;

23. Cao W, Fang Z, Hou G, et al. The psychological impact of the COVID-19 epidemic on college students in China. Psychiatry Res 2020;

24. Walter LA, McGregor AJ. Sex-and Gender-specific Observations and Implications for COVID-19. West J Emerg Med 2020; :507–509.

25. Kisely S, Warren N, McMahon L, Dalais C, Henry I, Siskind D. Occurrence, prevention, and management of the psychological effects of emerging virus outbreaks on healthcare workers: rapid review and meta-analysis. BMJ 2020; 369.

26. Peterman A, Potts A, Donnell MO, et al. Working Paper 528 April 2020 Pandemics and Violence Against Women and Children. Cent Glob Dev 2020; :43.

27. Davies SE, Bennett B. A gendered human rights analysis of Ebola and Zika: Locating gender in global health emergencies. Int Aff 2016;

28. Mollona E, Aivazidou E, Barberio V, Cunico G, Pareschi L. A policy framework for tackling the economic and social impact of the COVID-19 crisis. 2019.

29. Spinelli M, Lionetti F, Pastore M, Fasolo M. Parents’ Stress and Children’s Psychological Problems in Families Facing the COVID-19 Outbreak in Italy. Front Psychol 2020;

30. Wang G, Zhang Y, Zhao J, Zhang J, Jiang F. Mitigate the effects of home confinement on children during the COVID-19 outbreak. Lancet 2020; 395:945– 947.

31. Kang SJ, Jung SI. Age-Related Morbidity and Mortality among Patients with COVID-19. Infect Chemother 2020;

32. Armitage R, Nellums LB. COVID-19 and the consequences of isolating the elderly. Lancet Public Heal 2020; 5:e256. Available at: http://dx.doi.org/10.1016/S2468-2667(20)30061-X.

33. Petretto DR, Pili R. Ageing and COVID-19 : What is the Role for Elderly People? Geriatrics 2020; 5:25.

34. Cohen O, Geva D, Lahad M, et al. Community Resilience throughout the Lifespan the Potential Contribution of Healthy Elders. PLoS One 2016; 11:1–14.

35. Meng H, Xu Y, Dai J, Zhang Y, Liu B, Yang H. Analyze the psychological impact of COVID-19 among the elderly population in China and make corresponding suggestions. Psychiatry Res. 2020;

36. Bobes-Bascarán T, Sáiz PA, Velasco A, et al. Early psychological correlates associated with COVID-19 in a Spanish older adult sample. Am J Geriatr Psychiatry 2020; :1–12. Available at: http://www.sciencedirect.com/science/article/pii/S1064748120304759.

37. Fernández-Aguilar L, Ricarte J, Ros L, Latorre JM. Emotional differences in young and older adults: Films as mood induction procedure. Front Psychol 2018;

38. Beck F, Léger D, Fressard L, Peretti-Watel P, Verger P. Covid-19 health crisis and lockdown associated with high level of sleep complaints and hypnotic uptake at the population level. J. Sleep Res. 2020;

39. Shanahan L, Steinhoff A, Bechtiger L, et al. Emotional Distress in Young Adults During the COVID-19 Pandemic: Evidence of Risk and Resilience From a Longitudinal Cohort Study. Psychol Med 2020;

40. Commodari E, La Rosa VL. Adolescents in quarantine during COVID-19 pandemic in Italy: Perceived health risk, beliefs, psychological experiences and expectations for the future. Front Psychol 2020; 11:2480.

41. Suzuki Y, Maeda N, Hirado D, Shirakawa T, Urabe Y. Physical activity changes and its risk factors among community-dwelling japanese older adults during the COVID-19 epidemic: Associations with subjective well-being and health-related quality of life. Int J Environ Res Public Health 2020; 17:1–12.

42. Maugeri G, Castrogiovanni P, Battaglia G, et al. The impact of physical activity on psychological health during Covid-19 pandemic in Italy. Heliyon 2020;

43. Rossi R, Socci V, Pacitti F, et al. Mental Health Outcomes Among Frontline and Second-Line Health Care Workers During the Coronavirus Disease 2019 (COVID-19) Pandemic in Italy. JAMA Netw open 2020; 3:e2010185. Available at: http://www.ncbi.nlm.nih.gov/pubmed/32463467.

44. Xiao J, Fang M, Chen Q, He B. SARS, MERS and COVID-19 among healthcare workers: A narrative review. J Infect Public Health 2020; 13:843–848. Available at: https://doi.org/10.1016/j.jiph.2020.05.019.

45. Simione L, Gnagnarella C. Differences Between Health Workers and General Population in Risk Perception, Behaviors, and Psychological Distress Related to COVID-19 Spread in Italy. Front Psychol 2020; 11.

46. Pappa S, Ntella V, Giannakas T, Giannakoulis VG, Papoutsi E, Katsaounou P. Prevalence of depression, anxiety, and insomnia among healthcare workers during the COVID-19 pandemic: A systematic review and meta-analysis. Brain. Behav. Immun. 2020;

47. Alonso J, Vilagut G, Mortier P, et al. Mental health impact of the first wave of COVID-19 pandemic on Spanish healthcare workers: A large cross-sectional survey. Rev Psiquiatr Salud Ment 2021;

48. Mortier P, Vilagut G, Ferrer M, et al. Thirty-day suicidal thoughts and behaviors among hospital workers during the first wave of the Spain COVID-19 outbreak. Depress Anxiety 2021;

49. Ramaci T, Barattucci M, Ledda C, Rapisarda V. Social stigma during COVID-19 and its impact on HCWs outcomes. Sustain 2020;

50. Robert R, Kentish-Barnes N, Boyer A, Laurent A, Azoulay E, Reignier J. Ethical dilemmas due to the Covid-19 pandemic. Ann. Intensive Care. 2020;

51. Zanin M, Xiao C, Liang T, et al. The public health response to the COVID-19 outbreak in mainland China: a narrative review. J Thorac Dis 2020;

52. Verweij M, van de Vathorst S, Schermer M, Willems D, de Vries M. Ethical Advice for an Intensive Care Triage Protocol in the COVID-19 Pandemic: Lessons Learned from The Netherlands. Public Health Ethics 2020;

53. Vincent JL, Creteur J. Ethical aspects of the COVID-19 crisis: How to deal with an overwhelming shortage of acute beds. Eur Hear journal Acute Cardiovasc care 2020;

54. Owens IT. Supporting nurses’ mental health during the pandemic. Nursing (Lond) 2020; 50:54–57.

55. Martínez-López JÁ, Lázaro-Pérez C, Gómez-Galán J, Fernández-Martínez M del M. Psychological Impact of COVID-19 Emergency on Health Professionals: Burnout Incidence at the Most Critical Period in Spain. J Clin Med 2020;

56. Nowicki GJ, Ślusarska B, Tucholska K, Naylor K, Chrzan-Rodak A, Niedorys B. The severity of traumatic stress associated with Covid-19 pandemic, perception of support, sense of security, and sense of meaning in life among nurses: Research protocol and preliminary results from poland. Int J Environ Res Public Health 2020;

57. Delgado D, Quintana FW, Perez G, et al. Personal safety during the Covid-19 pandemic: Realities and perspectives of healthcare workers in latin America. Int J Environ Res Public Health 2020;

58. Epstein EG, Delgado S. Understanding and addressing moral distress. Online J Issues Nurs 2010;

59. Tomlin J, Dalgleish-Warburton B, Lamph G. Psychosocial Support for Healthcare Workers During the COVID-19 Pandemic. Front Psychol 2020;

60. Heath C, Sommerfield A, von Ungern-Sternberg BS. Resilience strategies to manage psychological distress among healthcare workers during the COVID-19 pandemic: a narrative review. Anaesthesia. 2020;

61. Johns Hopkins Berman Institute of Bioethics. Resources for Frontline Clinicians. 2020.

62. Maves RC, Downar J, Dichter JR, et al. Triage of Scarce Critical Care Resources in COVID-19 An Implementation Guide for Regional Allocation: An Expert Panel Report of the Task Force for Mass Critical Care and the American College of Chest Physicians. Chest. 2020;

