## Supplementary material for "Psychosocial impact of the Covid-19 pandemic: Identification of most vulnerable populations": Survey Questionnaire

| QUESTIONS | SCORING CODE |
|---|---|
| **General demography** | |
| How old are you? | |
| Which gender do you identify with? | male, female, non binary, I prefer not to say |
| In which country do you live? | |
| In which postal code do you live? | |
| How would you define your civil status? | single, married, divorced, widow, in a couple |
| Where do you live? | my own house/apartment, shared house/apartment, in a rented room, institutionalized, I am homeless |
| What level of education do you have? (check the maximum obtained) | primary education, secondary education, further education, bachelor degree, masters degree, doctoral degree |
| What is your job? | skilled job, unskilled job, caring for others/home, I have a company, I am self-employed, I am a healthcare worker (or working in a healthcare setting, I am unemployed, others |
| **Questions for the Scale of socio-economic precariousness** | For index scoring, sum of all points multiplied by 2. |
| Who provides financially at home? | >2 of us = 0 p, 2 of us =1 p, only me = 2p |
| Have you lost your job due to the COVID-19 outbreak? | no= 0 p; yes, the company made a temporary labour force adjustment plan= 1 p; yes, others = 1.5 p; yes, I was fired/the company made a labour force adjustment plan/ I have lost some jobs previously contracted/arranged = 2 p |
| Do you have savings? | yes= 0 p, yes, some= 1 p, no = 2 p |
| Do you have a mortgage to pay? | no = 0 p; yes, one =1 p; yes, more than 1 = 2 p |
| Do you have rent to pay? | no = 0 p, yes =2 p |
| Are you spending less since the COVID-19 outbreak? | no = 0 p; a little = 1 p; yes =2 p |
| Have you asked for social assistance or for any other assistance due to the COVID-19 outbreak? | no = 0 p; no, but will have to = 1 p; yes =2 p |
| Do you have to take care of somebody? (multiple choice question) | no = 0 p; yes (any answer: children <16 y.o., >16 y.o, parents, siblings, others) = 1 p per positive answer. |
| **Habits and COVID-19-related health status during confinement** | |
| (If having children): In which grade do you think the confinement is being difficult for children (and therefore for the family? | scale of potential answer, 0 being= not at all and 10= a lot |
| Are you staying at home, during this time? | yes; yes, I am teleworking; no, I work in essential services; no, I need to work; no, my employer does not allow me to |
| Are you scared or worried? | no; yes, of getting infected; yes, of going to the shops; yes, of infecting others; yes, that people close to me get infected |
| Who are you scared of infecting? | the children; my parents/close elderly people; my colleagues; anyone |
| Do you think you are consuming more since the outbreak began? | no; yes, I eat more; yes, I drink more (alcoholic drinks); yes, I smoke more; yes, I consume more illegal drugs; yes, I consume more drugs to calm myself down (sleeping pills, muscle relaxants, tranquilizers) |
| Through which channel do you receive information about the outbreak? | TV; Radio; Newspaper; Social media (Whatsapp, Twitter, Telegram etc.); Other channels |
| What do you think of the information you are receiving? | It's too much: I would like the Government to explain less; It's too much: I would like the media to explain less; It's too little : I would like the Government to explain more; It's too little : I would like the media to explain more; It's too negative/too sensationalist; I think it's poorly adjusted to reality; It's alright; I do not think anything about it |
| Do you think this situation has changed you? | no; yes, my life has changed; yes, my personality had changed; yes, the way I see society/the way we lived |
| Have you been in contact with someone infected by SARS-CoV-2? | yes, with a confirmed case (test positive); yes, with a probable non-confirmed case (test negative or test not done); I do not know |
| Since February, have you had any of these symptoms? | no; persistent cough (for one week or more); headache; persistent fever (for one week or more); extreme fatigue/tiredness; sore throat; muscle pain; loss of appetite/weight; loss of smell, smell blindness; loss of taste; diarrhea; dizziness; shortness of breath; chest pain; nasal congestion/running nose |
| How do you feel now? | well, normal, I do not feel at 100%, bad |
| In the last 14 days, have you used any healthcare resources put in place for the COVID-19 pandemic? | have called a telephone number set up for the management of COVID cases; have gone to the emergency room; have used an app set up for management of COVID cases; have been to a public healthcare center (including GP); have been to private doctor/healthcare center; have been tested; none of the above |
| If you were tested, what was the result? | positive, negative |
| **For HealthCare workers** | |
| What is your job? | physician, nurse, nurse assitant, technician, caretakr, researcher, kitchen personnel, cleaning personnel, administrative personnel, others |
| Have you been working with COVID patients directly? | no; not as far as I know; yes, I have been/am in a COVID team; yes, on duty |
| Are you scared of working with COVID patients? | no; yes, o being infected; yes, of dying; yes, of transmitting the virus to other non-COVID patients; yes, of transmitting the virus to my people (family/colleagues): yes, of being obliged to take medical decisions representing an ethical dilemma for me (patient selection, applicaton of protocols) |
| Have you had ethical concerns while working? | no; no, I think I need to follow the protocols; yes, with selection of patients and/or protocols for selection of patients or therapeutic indications; yes, others |
| **Questions related to mental-health** | **Scoring** |
| **Questions related to anxiety- How these sentences apply to you?** | For each of the questions below: never = 0 p, sometimes = 1 p, often = 2 p, almost always = 3 p. For the index scoring, sum of all points multiplied by 2. |
| last week I was aware of dryness of my mouth | |
| last week I experienced breathing difficulty (excessively rapid breathing, breathlessness in the absence of any physical exertion and absence of any | |
| last week I experienced trembling (eg in the hands) | |
| last week I was worried about situations in which I might panic ad make a fool of myself | |
| last week I felt I was close to panic | |
| last week I was aware of the action of my heart in the absence of physical exertions (sense of heart rate increase, heart missing a beat) | |
| last week I felt scared without any good reason | |
| **Questions related to stress- How these sentences apply to you?** | For each of the questions below: never = 0 p, sometimes = 1 p, often = 2 p, almost always = 3 p. For the index scoring, sum of all points multiplied by 2. |
| last week I found it hard to wind down | |
| last week I tended to over-react to situations | |
| last week I felt that I was using a lot of nervous energy | |
| last week I found myself getting agitated | |
| last week I found it difficult to relax | |
| last week I was intolerant of anything that kept me from getting on with what I was doing | |
| last week I felt that I was rather touchy | |
| **Questions related to depression- How these sentences apply to you?** | For each of the questions below: never = 0 p, sometimes = 1 p, often = 2 p, almost always = 3 p. For the index scoring, sum of all points multiplied by 2. |
| last week I couldn't seem to experience any positive feeling at all | |
| last week I found it difficult to work up the initiative to do things | |
| last week I felt that I had nothing to look forward to | |
| last week I felt down-hearted and blue | |
| last week I was unable to become enthusiastic about anything | |
| last week I felt that life was meaningless | |
| **Questions related to PSTD symptoms- How these sentences apply to you?** | For each of the questions below: 0= not at all, 1= a little bit, 2= moderately, 3= quite a bit, 4=extremely. For the index scoring, sum of all points multiplied by 2. |
| **Questions related to Intrusion symptoms** | |
| last week any reminder brought back feelings about it | |
| last week I had trouble staying asleep | |
| last week other things kept making me think about it. | |
| last week I thought about it when I didn't mean to | |
| last week Pictures about it popped into my mind | |
| last week I found myself acting or feeling like I was back at that time | |
| last week I had waves of strong feelings about it | |
| **Questions related to Avoidance symptoms** | |
| last week I avoided letting myself get upset when I thought about it or was reminded of it | |
| last week I felt as if it hadn't happened or wasn't real | |
| last week I stayed away from reminders of it. | |
| last week I thought about it when I didn't mean to | |
| last week I was aware that I still had a lot of feelings about it, but I didn't deal with them | |
| last week My feelings about it were kind of numb | |
| last week I tried to remove it from my memory | |
| last week I tried not to talk about it | |
| **Questions related to Hyperarousal symptoms** | |
| last week I felt irritable and angry | |
| last week I was jumpy and easily startled | |
| last week I had trouble falling asleep | |
| last week I had trouble concentrating | |
| last week I felt watchful and on-guard | |
