## Supplementary material for "Psychosocial impact of the Covid-19 pandemic: Identification of most vulnerable populations": Results broken down by category and according to gender and age group

| ANSWER CATEGORIES | | Conditional distributions given the responders gender (%) | | | | p (women vs men) | Conditional distributions given the responders age range (%) | | | | p |
|---|---|---|---|---|---|---|---|---|---|---|---|
| | | women | men | non binary | not saying | | <42 y.o. | 42- 52 y.o. | 52- 61 y.o. | >61 y.o. | |
| **Civil status** | Married | 51.04 | 60.21 | 14.77 | 27.94 | | 32.2 | 56.74 | 61.12 | 63.22 | |
| | Divorced | 11.75 | 7.94 | 5.68 | 16.17 | | 2.52 | 11.33 | 15.14 | 13.08 | |
| | In couple | 18.49 | 17.39 | 39.77 | 23.52 | | 38.02 | 18.34 | 10.91 | 6.85 | |
| | Single | 14.51 | 12.89 | 38.63 | 30.88 | | 27.15 | 12.85 | 10.06 | 7.2 | |
| | Widow | 4.18 | 1.54 | 1.13 | 1.47 | | 0.08 | 0.71 | 2.75 | 9.63 | |
| **Housing** | Owned appartment/house | 91.08 | 90.89 | 64.36 | 72.46 | | 79.44 | 94.22 | 95.08 | 94.48 | p<0.01 |
| | Shared appartment/house | 7.7 | 7.9 | 26.43 | 23.18 | | 18.43 | 4.9 | 4.11 | 4.33 | |
| | Rented room | 1.05 | 1.07 | 8.04 | 0,00 | | 2.01 | 0.81 | 0.67 | 0.83 | |
| | Centre/institution | 0.13 | 0.09 | 0,00 | 0,00 | | 0.05 | 0.03 | 0.09 | 0.3 | |
| | Homeless | 0.02 | 0.03 | 1.14 | 4.34 | | 0.05 | 0.01 | 0.02 | 0.03 | |
| **Maximum Education Degree** | Primary Education | 3.52 | 4.63 | 5.68 | 5.79 | p<0.01 | 1.53 | 3.3 | 4.24 | 6.1 | p<0.01 |
| | Secondary Education | 5.18 | 6.17 | 3.4 | 1.44 | | 4.83 | 4.49 | 5.19 | 7.19 | |
| | High School | 29.92 | 35.46 | 29.54 | 28.98 | | 27.54 | 30.98 | 34.17 | 33.11 | |
| | Degree | 44.99 | 36.96 | 31.81 | 33.33 | | 38.72 | 43.92 | 43.48 | 44.26 | |
| | Master | 13.47 | 12.77 | 26.13 | 21.73 | | 24.32 | 14.3 | 9.7 | 5.65 | |
| | PhD | 2.9 | 3.98 | 3.4 | 8.69 | | 3.03 | 2.99 | 3.2 | 3.67 | |
| **Employment** | Qualified job | 36.95 | 34.15 | 35.22 | 37.68 | p<0.01 | 48.19 | 48.76 | 41.3 | 7.86 | 0<0.01 |
| | Non qualified job | 3.51 | 3.78 | 9.09 | 2.89 | | 4.39 | 4.46 | 4.49 | 1.15 | |
| | Job in Healthcare | 10.9 | 4.67 | 9.09 | 1.44 | | 12.16 | 10.58 | 9.21 | 4.64 | |
| | Home/people care | 6.24 | 1.42 | 0,00 | 2.89 | | 0.94 | 1.69 | 3.25 | 12.86 | |
| | Self-employed | 8.03 | 11.41 | 9.09 | 15.94 | | 7.72 | 11.45 | 11.4 | 5.59 | |
| | Company owner | 3,00 | 7.36 | 1.13 | 1.44 | | 2.39 | 5.66 | 5.9 | 3.05 | |
| | Unemployed | 5.29 | 4.54 | 12.5 | 11.59 | | 7.63 | 4.62 | 5.61 | 2.69 | |
| | Other | 26.03 | 32.63 | 23.86 | 26.08 | | 16.54 | 12.73 | 18.8 | 62.13 | |
| **People financially providing at home** | >2 | 8.03 | 7.05 | 14.77 | 16.41 | | 13.59 | 3.99 | 7.26 | 6.43 | |
| | 2 | 66.29 | 68.57 | 54.54 | 55.22 | | 70.39 | 71.65 | 64.94 | 61.18 | |
| | 1 | 25.67 | 24.37 | 30.68 | 28.35 | | 16012,00 | 24.35 | 27.78 | 32.38 | |
| **Care of someone** | No | 36.55 | 47.61 | 58.94 | 34.66 | <0.01 | 45.98 | 16.26 | 31.39 | 67.82 | <0.01 |
| | Yes, of people of <16 y.o. | 25.99 | 21.93 | 13.68 | 25.33 | | 33.96 | 48.69 | 13.52 | 3.07 | |
| | Yes, of people of >16 y.o. | 13.02 | 10.35 | 6.31 | 6.66 | | 4.81 | 12.58 | 23.54 | 6.73 | |
| | Yes, siblings | 1.36 | 0.96 | 4.21 | 2.66 | | 1.57 | 0.86 | 1.33 | 1.28 | |
| | Yes, parents | 16.1 | 12.66 | 10.52 | 17.33 | | 8.41 | 16.92 | 23.03 | 10.92 | |
| | Yes, others | 6.95 | 6.46 | 6.31 | 13.33 | | 5.24 | 4.66 | 7.17 | 10.16 | |
| **Burden of care** | None | 40.62 | 51.34 | | | <0.01 | 48.85 | 18.85 | 35.96 | 70.47 | <0.01 |
| | 1 option selected | 25.9 | 22.11 | | | | 13.12 | 21.03 | 39.98 | 24.24 | |
| | 2 options selected | 28.23 | 22.83 | | | | 34.82 | 49.43 | 19.32 | 4.51 | |
| | 3 options selected | 4.77 | 3.39 | | | | 2.82 | 9.88 | 4.31 | 0.61 | |
| | 4 options selected | 0.41 | 0.30 | | | | 0.31 | 0.73 | 0.38 | 0.11 | |
| | 5 options selected | 0.04 | 0.01 | | | | 0.04 | 0.04 | 0.02 | 0.02 | |
| **Loss of job** | No | 76.13 | 72.73 | 63.63 | 65.21 | <0.01 | 68.4 | 69.41 | 73.65 | 88.18 | <0.01 |
| | Yes, the company made a labour force adjustment plan | 0.18 | 0.17 | 0,00 | 0,00 | | 0.22 | 0.26 | 0.15 | 0.09 | |
| | Yes, the company made a temporary labour force adjustment plan | 9.70 | 10.01 | 9.09 | 7.24 | | 14.5 | 13.04 | 9.9 | 2.17 | |
| | Yes, I have lost some jobs previously contracted/arranged | 4.93 | 7.61 | 15.9 | 14.49 | | 6.75 | 7.17 | 6.68 | 2.54 | |
| | Yes, I was fired | 0.96 | 0.68 | 2.27 | 0,00 | | 1.79 | 0.96 | 0.67 | 0.16 | |
| | Yes, others | 8.08 | 8.77 | 9.09 | 13.04 | | 8.3 | 9.12 | 8.93 | 6.83 | |
| **Savings** | No | 22.00 | 18.00 | 30,00 | 26,00 | <0.01 | 20.34 | 24.48 | 22.21 | 15.82 | <0.01 |
| | Yes | 34.00 | 40.00 | 23,00 | 28,00 | | 36.22 | 32.37 | 33.65 | 40.55 | |
| | Some | 44.00 | 42.00 | 48,00 | 46,00 | | 43.43 | 43.14 | 44.13 | 43.62 | |
| **Mortgage to pay** | No | 58.75 | 59.47 | 80.68 | 57.97 | <0.01 | 64.04 | 39.65 | 54.68 | 76.91 | <0.01 |
| | Yes, one | 36.17 | 34.37 | 18.18 | 36.23 | | 31.76 | 50.8 | 39.81 | 20.66 | |
| | Yes, more than one | 5.07 | 6.14 | 1.13 | 5.79 | | 4.18 | 9.54 | 5.49 | 2.42 | |
| **Rent to pay** | No | 76.00 | 76.00 | 51,00 | 66,00 | | 56.64 | 75.05 | 83.23 | 87.08 | <0.01 |
| | Yes | 24.00 | 24.00 | 49,00 | 34,00 | | 43.35 | 24.94 | 16.76 | 12.91 | |
| **Spending less** | Yes | 59.85 | 62.61 | 59.09 | 69.56 | | 64.15 | 58.86 | 60.4 | 59.52 | |
| | A little | 22.34 | 21.56 | 13.63 | 17.39 | | 19.89 | 23.74 | 22.72 | 21.87 | |
| | No | 17.80 | 15.82 | 27.27 | 13.04 | | 15.95 | 17.38 | 16.87 | 18.59 | |
| **Seek for social assistance/or any other assistance** | No | 91.42 | 90.8 | 80.68 | 81.15 | | 88.95 | 88.41 | 90.73 | 96.48 | <0.01 |
| | Not yet, but will need to | 4.71 | 5.19 | 10.22 | 8.69 | | 6.34 | 6.43 | 5.08 | 1.81 | |
| | Yes | 3.85 | 3.99 | 9.09 | 10.14 | | 4.7 | 5.15 | 4.18 | 1.7 | |
| **Index of socio-economic deprivation -score** | <7 | 26.19 | 17.04 | 22.47 | 17.39 | <0.01 | 21.17 | 30.35 | 26.04 | 22.72 | p<0.01 |
| | 7-8.5 | 20,00 | 10.22 | 20.12 | 10.14 | | 33.2 | 28.42 | 32.07 | 36.36 | |
| | 8.5-10 | 32.09 | 32.95 | 33.59 | 43.47 | | 17.38 | 18.8 | 19.27 | 24.3 | |
| | >10 | 21.71 | 39.77 | 23.8 | 28.98 | | 28.24 | 22.41 | 22.6 | 16.59 | |
| **Staying home** | No, I am forced to go to work | 0.33 | 0.55 | 2.29 | 1.44 | <0.01 | 0.54 | 0.56 | 0.4 | 0.1 | p<0.01 |
| | No, I need to work | 0.69 | 1.51 | 1.14 | 1.44 | | 0.75 | 0.79 | 0.88 | 1.3 | |
| | No, I work on essential services | 13.73 | 12.39 | 13.79 | 7.24 | | 16.36 | 17.77 | 15.19 | 4.47 | |
| | Yes | 54.13 | 57.73 | 43.67 | 62.31 | | 43.85 | 39.51 | 48.13 | 87.39 | |
| | Yes, teleworking | 31.1 | 27.79 | 39.08 | 27.53 | | 38.48 | 41.35 | 35.37 | 6.71 | |
| **Afraid** | No | 22.14 | 38.44 | 26.26 | 37.68 | <0.01 | 21.77 | 23.06 | 26.82 | 35.04 | p<0.01 |
| | Yes, going shopping | 18.9 | 13.39 | 17.17 | 10.14 | | 17.82 | 18.59 | 16.69 | 16.19 | |
| | Yes, to infect others | 23.89 | 17.68 | 30.3 | 24.63 | | 28.52 | 24.76 | 22.13 | 13.85 | |
| | Yes, to get infected | 35.04 | 30.47 | 26.26 | 27.53 | | 31.87 | 33.57 | 34.33 | 34.9 | |
| **Afraid to infect** | Elders | 36.23 | 34.25 | 43.33 | 23.52 | | 42.05 | 35.33 | 36.86 | 22.98 | p<0,01 |
| | Anyone | 48.63 | 51.26 | 50,00 | 70.58 | | 41.27 | 41.49 | 54.17 | 69.79 | |
| | Children | 13.32 | 12.97 | 3.33 | 5.88 | | 14.28 | 21.55 | 7.21 | 6.47 | |
| | Colleagues at work | 1.81 | 1.50 | 3.33 | 0,00 | | 2.38 | 1.61 | 1.74 | 0.74 | |
| **Increased consume of substances** | No | 55.2 | 64.77 | 41.22 | 50,00 | <0.01 | 42.95 | 51.97 | 59.86 | 77.68 | p<0.01 |
| | Yes, alcohol | 5.57 | 6.74 | 8.77 | 9.75 | | 8.88 | 7.23 | 5.01 | 2.47 | |
| | Yes, food | 26.26 | 19.40 | 22.8 | 20.73 | | 33.04 | 27.44 | 22.72 | 13.4 | |
| | Yes, illegal drugs | 0.25 | 0.73 | 5.26 | 2.43 | | 1.07 | 0.28 | 0.16 | 0.09 | |
| | Yes, drugs to calm down | 4.83 | 2.44 | 8.77 | 6.09 | | 4.24 | 4.99 | 4.27 | 3.07 | |
| | Yes, tobacco | 7.85 | 5.89 | 13.15 | 10.97 | | 9.79 | 8.06 | 7.95 | 3.27 | |
| **Media to get information about the pandemic** | Social media | 30.09 | 27.20 | 35,00 | 30.88 | | 7.49 | 5.45 | 3.41 | 1.49 | |
| | TV | 37.48 | 35.18 | 28.33 | 31.61 | | 50.54 | 50.41 | 50083,00 | 48.38 | |
| | Radio | 14.94 | 16.67 | 10,00 | 12.5 | | 13.74 | 20.14 | 22.9 | 25.1 | |
| | Newspapers | 12.83 | 15.18 | 15,00 | 11.76 | | 19.17 | 16.7 | 17.07 | 20.19 | |
| | Other | 4.63 | 5.74 | 11.66 | 13.23 | | 9.03 | 7.27 | 6.52 | 4.82 | |
| **Thoughts about the information received** | It's ok | 19.28 | 18.40 | 6.33 | 13.18 | <0.01 | 9.76 | 17.8 | 28.13 | 26.74 | <0.01 |
| | The Government explains too much | 2.65 | 4.55 | 0,00 | 2.19 | | 1.44 | 2.28 | 3.88 | 6.66 | |
| | The Government explains too less | 9.06 | 8.60 | 14.08 | 9.89 | | 8.99 | 8.56 | 9.7 | 8.53 | |
| | Media explain too much | 12.49 | 13.43 | 11.97 | 8.79 | | 9.69 | 10.46 | 14.32 | 19.21 | |
| | Media explain too less | 2.8 | 3.11 | 5.63 | 8.79 | | 2.68 | 2.69 | 3.53 | 2.96 | |
| | Too negative | 20.47 | 21.90 | 25.35 | 18.68 | | 41.88 | 26.09 | 0.24 | 0.11 | |
| | Poorly adjusted to the reality | 27.34 | 25.60 | 30.98 | 29.67 | | 21.13 | 25.61 | 33.57 | 31.12 | |
| | I do not think anything about it | 5.87 | 4.36 | 5.63 | 8.79 | | 4.38 | 6.47 | 6.6 | 4.64 | |
| **Impact of the pandemic on people (subjective)** | No | 18.11 | 29.88 | 23.07 | 23.25 | <0.01 | 17.23 | 19.43 | 21.13 | 28.05 | <0.01 |
| | Yes, my personality | 5.18 | 3.71 | 9.4 | 5.81 | | 8.17 | 5.55 | 3.29 | 2.02 | |
| | Yes, my vision of the society/ ho | 51.74 | 47.05 | 43.58 | 50,00 | | 50.98 | 51.86 | 52.4 | 46.36 | |
| | Yes, my life | 24.95 | 19.34 | 23.93 | 20.93 | | 23.6 | 23.14 | 23.17 | 23.56 | |
| **Contact with someone infected by SARS-CoV-2** | I do not know | 79.01 | 82.93 | 70.32 | 82.6 | <0.01 | 75,00 | 76.77 | 79.62 | 88.72 | <0.01 |
| | yes, with a probable non-confirmed case | 10.16 | 9.01 | 16.48 | 5.79 | | 13.05 | 11.61 | 9.79 | 5.14 | |
| | Yes, with a confirmed case | 10.81 | 8.04 | 13.18 | 11.59 | | 11.93 | 11.61 | 10.58 | 6.12 | |
| **Presence of symptoms (since February)** | No | 22.92 | 35.72 | 11.29 | 37.75 | | 15.55 | 20.98 | 28.09 | 46.06 | |
| | Headache | 17.06 | 13.02 | 13.7 | 8.16 | | 17.59 | 18.01 | 16.29 | 10.81 | |
| | Sore throat | 10.51 | 7.95 | 9.27 | 13.26 | | 10.81 | 10.59 | 9.47 | 7.96 | |
| | Nasal congestion/running nose | 9.1 | 9.37 | 10.08 | 12.24 | | 12.06 | 9.05 | 8.28 | 6.2 | |
| | Extreme fatigue/tiredness | 7.47 | 5.30 | 10.48 | 4.08 | | 7.92 | 7.57 | 6.76 | 4.77 | |
| | Persistent cough (for one week or | 6.96 | 6.50 | 6.85 | 7.14 | | 6.71 | 6.94 | 6.92 | 6.81 | |
| | Muscle pain | 6.55 | 5.15 | 8.87 | 4.08 | | 6.54 | 6.78 | 6.43 | 4.67 | |
| | Diarrhea | 5.37 | 5.32 | 8.46 | 6.12 | | 6.74 | 5.63 | 5.06 | 3.36 | |
| | Dizziness | 3.14 | 1.95 | 8.06 | 2.04 | | 3.92 | 2.97 | 2.53 | 1.54 | |
| | Shortness of breath | 2.27 | 1.95 | 3.62 | 2.04 | | 2.88 | 2.48 | 1.88 | 1.19 | |
| | Chest pain | 1.96 | 1.74 | 1.2 | 2.04 | | 2.38 | 2.28 | 1.71 | 0.93 | |
| | Loss of smell, smell blindness | 1.93 | 1.66 | 2.41 | 1.02 | | 2.15 | 2.05 | 1.76 | 1.31 | |
| | Persistent fever (for one week or more) | 1.58 | 1.79 | 2.41 | 0,00 | | 1.5 | 1.5 | 1.83 | 1.76 | |
| | Loss of appetite/weight | 1.38 | 1.10 | 2.01 | 0,00 | | 1.38 | 1.3 | 1.26 | 1.28 | |
| | Loss of taste | 1.74 | 1.42 | 1.2 | 0,00 | | 1.79 | 1.79 | 1.66 | 1.28 | |
| **How did they feel when answering the questionnaire** | Well | 64.92 | 70.28 | 52.87 | 60.86 | | 68.25 | 67.4 | 64.28 | 65.97 | <0.01 |
| | Normal | 22.84 | 21.6 | 18.39 | 24.63 | | 19.31 | 19.85 | 23.86 | 26.65 | |
| | Not at 100% | 11.76 | 7.83 | 25.28 | 13.04 | | 11.93 | 12.26 | 11.39 | 7.13 | |
| | Bad | 0.46 | 0.27 | 3.44 | 1.44 | | 0.5 | 0.46 | 0.45 | 0.23 | |
| **Use of healthcare resources put in place in the context of the COVID-19 pandemic** | None | 63.97 | 64.91 | 55.33 | 78.57 | <0.01 | 63.21 | 62.37 | 62.73 | 68.55 | <0.01 |
| | Have used an app set up for management of COVID cases | 20.9 | 22.99 | 26.21 | 17.14 | | 20.53 | 22.12 | 23.33 | 20.05 | |
| | Have called a telephone number set up for the management of COVID cases | 5.9 | 4.89 | 4.85 | 1.42 | | 6.56 | 6.59 | 5.49 | 3.83 | |
| | Have been to a public healthcare center (including GP) | 3.97 | 3.27 | 2.91 | 1.42 | | 3.97 | 3.96 | 3.56 | 3.58 | |
| | Have been tested | 2.1 | 1.14 | 3.88 | 1.42 | | 2.33 | 2.06 | 1.85 | 1.07 | |
| | Have been to private doctor/healthcare center | 1.69 | 1.39 | 1.94 | 0,00 | | 1.76 | 1.36 | 1.48 | 1.82 | |
| | Have gone to the emergency room | 1.43 | 1.37 | 4.85 | 0,00 | | 1.6 | 1.5 | 1.53 | 1.08 | |
| **For those tested, result of the test** | Negative | 61.14 | 42.48 | 50,00 | 100,00 | | 62.05 | 59.21 | 54.25 | 51.7 | |
| | Positive | 38.85 | 57.51 | 50,00 | 0,00 | | 37.94 | 40.78 | 45.74 | 48.29 | |
| **Occupation** | Nurse | 33,70 | 13,62 | 25,00 | 0,00 | <0.01 | 34,33 | 29,26 | 28,42 | 28,59 | <0.01 |
| | Physician | 17,96 | 42,67 | 12,50 | 0,00 | | 16,22 | 17,26 | 22,22 | 43,70 | |
| | Others (including working on a private pharmacy) | 12,74 | 13,88 | 0,00 | 0,00 | | 15,98 | 13,71 | 10,58 | 8,15 | |
| | Technician | 11,19 | 13,36 | 0,00 | 0,00 | | 11,97 | 13,85 | 10,81 | 6,67 | |
| | Administrative personnel | 10,66 | 6,23 | 25,00 | 0,00 | | 7,59 | 11,87 | 13,53 | 4,89 | |
| | Nurse assistant | 10,56 | 3,89 | 37,50 | 0,00 | | 9,54 | 9,82 | 11,26 | 6,07 | |
| | Researcher | 2,36 | 3,24 | 0,00 | 100,00 | | 3,22 | 2,80 | 1,89 | 1,48 | |
| | Caretaker | 0,18 | 2,59 | 0,00 | 0,00 | | 0,67 | 0,55 | 0,60 | 0,15 | |
| | Cleaning personnel | 0,28 | 0,39 | 0,00 | 0,00 | | 0,30 | 0,41 | 0,30 | 0,00 | |
| | Kitchen personnel | 0,28 | 0,13 | 0,00 | 0,00 | | 0,12 | 0,48 | 0,15 | 0,30 | |
| | Laundry personnel | 0,09 | 0,00 | 0,00 | 0,00 | | 0,06 | 0,00 | 0,23 | 0,00 | |
| **Having worked directly with COVID-19 patients** | No | 56,88 | 56,32 | 50,00 | 100,00 | <0.01 | 50,23 | 53,55 | 60,78 | 72,67 | <0.01 |
| | Yes | 43,12 | 43,68 | 50,00 | 0,00 | | 49,77 | 46,45 | 39,22 | 27,33 | |
| **Fear of working with COVID-19 patients** | No | 21,97 | 36,04 | | | <0.01 | 18,70 | 20,17 | 25,18 | 31,70 | <0.01 |
| | Yes | 78,03 | 63,96 | | | | 81,30 | 79,83 | 74,82 | 68,30 | |
| **Fear of working with COVID-19 patients** | No fear | 16,81 | 28,40 | 21,43 | 0,00 | <0.05 | 11,61 | 14,50 | 17,70 | 22,00 | <0.01 |
| | Scared of transmitting the virus to other non-COVID patients | 19,30 | 17,40 | 14,29 | 0,00 | | 17,30 | 16,22 | 13,93 | 12,93 | |
| | Scared of transmitting the virus to own people (family, colleagues) | 55,81 | 47,14 | 35,71 | 100,00 | | 46,58 | 45,97 | 44,23 | 38,32 | |
| | Scared of being obliged to take medical decisions representing an ethical dilemma for me (patient selection, application of protocols) | 8,09 | 7,07 | 14,29 | 0,00 | | 8,23 | 6,72 | 4,72 | 4,99 | |
| | Scared of being infected | 21,69 | 21,32 | 14,29 | 0,00 | | 16,27 | 16,59 | 19,42 | 21,77 | |
| | Afraid of dying | 5,43 | 5,50 | 7,14 | 0,00 | | 4,05 | 4,30 | 4,45 | 4,55 | |
